# Causal role of medial superior frontal cortex on enhancing neural information flow and self-agency judgments in the self-agency network

**DOI:** 10.1101/2024.02.13.24302764

**Authors:** Yingxin Jia, Kiwamu Kudo, Namasvi Jariwala, Phiroz Tarapore, Srikantan Nagarajan, Karuna Subramaniam

## Abstract

Self-agency is being aware of oneself as the agent of one’s thoughts and actions. Self-agency is necessary for successful interactions with the outside world (reality-monitoring). Prior research has shown that the medial superior prefrontal gyri (mPFC/SFG) may represent one neural correlate underlying self-agency judgments. However, the causal relationship remains unknown. Here, we applied high-frequency 10Hz repetitive transcranial magnetic stimulation (rTMS) to modulate the excitability of the mPFC/SFG site that we have previously shown to mediate self-agency. For the first time, we delineate *causal* neural mechanisms, revealing precisely how rTMS modulates SFG excitability and impacts *directional* neural information flow in the self-agency network by implementing innovative magnetoencephalography (MEG) phase-transfer entropy (PTE) metrics, measured from pre-to-post rTMS. We found that, compared to control rTMS, enhancing SFG excitability by rTMS induced significant increases in information flow between SFG and specific cingulate and paracentral regions in the self-agency network in delta-theta, alpha, and gamma bands, which predicted improved self-agency judgments. This is the first multimodal imaging study in which we implement MEG PTE metrics of 5D imaging of space, frequency and time, to provide cutting-edge analyses of the *causal* neural mechanisms of how rTMS enhances SFG excitability and improves neural information flow between distinct regions in the self-agency network to potentiate improved self-agency judgments. Our findings provide a novel perspective for investigating *causal* neural mechanisms underlying self-agency and create a path towards developing novel neuromodulation interventions to improve self-agency that will be particularly useful for patients with psychosis who exhibit severe impairments in self-agency.

## Introduction

Self-agency is the awareness of oneself as the agent of one’s thoughts and actions. Self-agency is of cardinal importance because it is integral to self-awareness in the context of human interactions with the outside world (i.e., reality monitoring) (1, 2). Reality monitoring is the ability to differentiate internal self-generated information from externally-derived information. In our prior reality monitoring studies, in which subjects distinguish self-generated from externally-derived information, healthy control participants (HC) showed increased medial superior prefrontal gyri (mPFC/SFG) activity during the successful encoding and retrieval of self-generated information, which correlated with accurate judgments of self-agency (i.e., accurate identification of self-generated information), indicating this mPFC/SFG site represents one neural correlate of self-agency (1, 3, 4).

The mPFC/SFG site represents a critical brain region that consistently exhibits heightened activity before self-generated actions (that do not occur prior to externally-perceived actions). This increased mPFC/SFG self-preparatory activity which is thought to lead to the experience of self-agency, has been observed across convergent imaging studies (functional MRI, magnetoencephalography (MEG), and electroencephalography (EEG)) as well as single neuron studies (3, 5–10). Given these correlative data that mPFC/SFG supports self-agency in HC, we now test whether enhancing SFG excitability can causally modulate self-agency during a reality-monitoring task.

In the present study, for the first time we use repetitive transcranial magnetic stimulation (rTMS) as a causal neurostimulation tool to test whether increasing SFG excitability with high-frequency 10Hz rTMS, will improve directed neural information flow in the self-agency network to enhance self-agency judgments during reality monitoring. Specifically, we implemented a double-blinded randomized controlled trial (RCT) in which HC are assigned to either active rTMS to enhance SFG excitability or 10Hz rTMS applied to a control distal site outside the self-agency network (N=15). To-date, the neural mechanisms as to how rTMS modulates the excitability of a targeted region, such as SFG, and induces neural plasticity via trans-synaptic propagation of neural information flow to its connected sites within the self-agency network, remain a mystery. The advent of innovative MEG phase-transfer entropy (PTE) metrics of 5-dimensional imaging of space, frequency and time, provide a unique opportunity to understand the *causal* neural mechanisms of how neurostimulation TMS techniques modulate not only neural activity in targeted TMS sites, but also induce neural plasticity, shown by *directional* neural information flow to connected regions in critical self-agency networks.

In this study, we capitalize on the powerful linkage of multimodal MRI/TMS/MEG that enables us to localize both spatial aspects of TMS with regard to each subject’s neuroanatomical MRI, and delineate with MEG the neurophysiological temporal and frequency band characteristics of SFG stimulation with respect to control TMS. Specifically, we use the high spatiotemporal resolution of MEG by implementing cutting-edge PTE analyses, measured from pre-to-post rTMS, which enable *causal* computations of the temporal propagation in *directed* neural information flow, measured not only in different MEG frequency spectra but with millisecond timing resolution, between the stimulated SFG region and other regions in the self-agency network that is induced by active rTMS to SFG, compared to baseline and control rTMS. In our prior studies, we have examined temporal correlations in resting-state neural oscillation which are quantified by the imaginary coherence between a voxel and the rest of the brain (11–13). Here, with MEG PTE analyses that are based on the principles of information theory, we are able to quantify how much information in the future of a region-of-interest (ROI) target is predictable when knowing the past state of the neural source (14–17). Thus, MEG PTE metrics enable us to move beyond correlative paradigms to provide causal computations of changes in directed neural information flow between brain regions, assayed from a neural source to a target, and enable the delineation of the precise mechanisms of trans-synaptic propagation of neural information flow within the self-agency network that are induced by rTMS to SFG, compared to rTMS of the control site (14–17). Taken together, here, we employ a causal multimodal imaging framework to delineate, for the first time, the precise neural mechanisms as to how enhancing medial SFG excitability with rTMS modulates the temporal propagation of directed neural information flow between distinct regions in the self-agency network to potentiate improved self-agency judgments.

We hypothesized that, compared to control rTMS, enhancing medial SFG excitability by rTMS while subjects are at rest would improve the temporal propagation of neural information flow from SFG to anatomically-connected distinct regions (such as the cingulate gyrus and premotor/supplementary motor cortex) within the self-agency network in either delta-theta (2-8 Hz), alpha (8-12 Hz), beta (12-30 Hz), or gamma (30-50 Hz) frequency bands. For example, we know that during the default mode, when participants are at rest and left to think to themselves, the lower frequency band oscillations such as delta-theta and alpha bands, which represent an idling rhythm, are dominant in medial frontal and adjacent cingulate cortex during rest across both MEG and EEG recordings, and functionally overlap with resting-state networks found in fMRI studies (18–21). We also know rTMS propagates trans-synaptically, and that SFG has direct connections with the cingulate gyrus (CG) and premotor/supplementary motor cortex (i.e., the paracentral lobule, PCL) in the self-agency network (22, 23). Further, convergent functional imaging studies across fMRI, PET and EEG studies, have shown increased neural activity in CG and PCL, immediately prior to subjects engaging in self-generated actions, compared to externally-triggered actions, which is thought to reflect subjects’ internal thoughts and the volition in motor preparation to translate these thoughts to initiate self-generated actions that results in the experience of self-agency (2, 9, 24, 25).

This neural activity increase in PCL is thought to mediate self-predictions in ‘self-generated forward models’ (also known as efference copies of the motor command sent to sensory cortices) that model the expected sensory outcome of self-generated actions, that lead to the experience of self-agency (26–28). The sensory outcome of self-generated actions is well-predicted, compared to external actions (29–31). Thus, typically the smaller the prediction error between the actual and predicted sensory feedback, the more likely the outcome will be attributed as a self-generated (29, 31, 32). For example, in our speech monitoring tasks, HC experience self-agency only when auditory sensory feedback (i.e., what we hear when we speak) slightly deviates from predictions of what they expect to hear (1, 31, 33). In these speech monitoring studies, we have found increased medial frontal activation extending to cingulate activation, shown in beta and gamma activity, during self-predictions of speech monitoring only when auditory feedback minimally deviates from predictions of what HC expect to hear, that results in the experience of self-agency (1, 34). Similarly, on a different reality-monitoring task, we also found increased medial frontal neural activity extending to cingulate activation, during the successful encoding and memory retrieval of self-generated information, which correlated with accurate judgments of self-agency (1, 3, 4). Overall, these findings suggest that medial frontal activation and the temporal propagation of neural connectivity and information flow to cingulate and paracentral lobule are critical nodes in the self-agency network that function to improve self-prediction mechanisms across disparate tasks that lead to a unitary experience of self-agency.

Taken these prior findings together, we hypothesized that compared to control rTMS, enhancing medial frontal excitability by rTMS would improve trans-synaptic neural plasticity in the self-agency network manifested as increased neural information flow from medial SFG to PCL and CG, and this improved neural information flow would predict improved self-agency judgments. If these hypotheses are confirmed, this would be the first study in which we implement state-of-the-art MEG PTE techniques, measured from pre-to post rTMS, to delineate the precise causal neural mechanisms underlying self-agency by revealing how enhancing SFG excitability by rTMS improves neural information flow between distinct regions in the self-agency network to potentiate improved self-agency judgments.

## Results

We found that compared to control rTMS, enhancing SFG excitability by rTMS induced significant improvement in self-agency judgments (Fig. 2A-B). To delineate the neural mechanisms underlying these self-agency improvements, we show for the first time that enhancing SFG excitability by rTMS increased information flow from SFG to distinct regions in the self-agency network (such as the cingulate gyrus and paracentral lobule) in delta-theta (2-8 Hz), alpha (8-12 Hz), beta (12-30 Hz), and gamma (30-50 Hz) frequency bands, which were significant at strict FDR thresholds, corrected for whole-brain multiple comparisons (FDR, p<.05) (see Figs. 3-6). Additionally, increased information flow from SFG to paracentral lobule (PCL) was observed in lower frequency bands such as delta-theta and alpha bands, whereas increased information flow from SFG to cingulate gyrus (CG) was observed in higher gamma frequencies, which predicted improved self-agency judgments on the reality-monitoring task only in the HC participants who completed rTMS targeting the SFG (see Figs. 3D, 4D 6C), but not in the control rTMS HC group (all p’s>.05). In other words, enhancing SFG excitability by rTMS increased information outflow from SFG to CG in all frequency bands; however, increase in information outflow to CG predicted better self-agency judgments in higher-frequency gamma bands while increase in information outflow from SFG to PCL predicted better self-agency judgments in lower-frequency delta-theta and alpha bands.

We did not find improved self-agency judgments on the reality-monitoring task or increased information flow in any regions after HC completed rTMS to the control left temporoparietal site, compared to baseline (see Supplementary Fig. 1) (all p’s >.05). Overall, our findings provide the first causal evidence to show that the improvements in self-agency judgments were specifically due to the effects of medial SFG stimulation, rather than due to non-specific neural changes in the control rTMS site, and thus cannot be attributed to general TMS effects. Taken together, the present MEG findings, quantified by PTE metrics, provide the high spatiotemporal resolution to accurately capture with millisecond precision the neural changes in information flow that are induced by rTMS to SFG, and delineate for the first time the causal neurophysiological mechanisms of SFG stimulation on modulating the self-agency network.

## Discussion

This study is the first comprehensive demonstration, revealing that rTMS applied to SFG not only induced targeted modulation of SFG excitability but also induced trans-synaptic neural plasticity by stimulating increased neural information flow from SFG to distinct sites in the self-agency network, such as PCL and CG, which predicted and potentiated improved self-agency judgements on the reality-monitory task. In particular, increased information flow from SFG to PCL was observed in lower frequency bands (i.e., delta-theta and alpha), whereas increased information flow from SFG to CG was found in higher gamma frequencies, which predicted improved self-agency judgments on the reality-monitoring task. Importantly, the findings of increased neural information flow between

SFG with PCL and CG that potentiated improved self-agency judgments, were only observed in the HC participants who completed rTMS targeting SFG but not in the control rTMS HC group. After HC completed control rTMS, we did not find any regions that showed increased neural information flow or improved self-agency judgments on the reality-monitoring task. Together, these convergent findings provide robust evidence that the increases in neural information flow between SFG and specific CG and PCL regions in the self-agency network, and their potentiation of improving self-agency judgments were specific to rTMS modulation of SFG excitability, and cannot be attributed to practice task effects or general effects of rTMS.

The medial superior frontal cortex extending to cingulate gyrus is a central hub within the default mode network (DMN), a network of brain regions that is particularly active when participants are left at rest to self-reflect on their inner thoughts (18–20, 25). In particular, lower frequency band oscillations such as delta-theta and alpha bands that are dominant in medial frontal cortex during resting-states, have been observed across convergent MEG, EEG and fMRI studies, and represent an idling rhythm associated with self-reflection and internal thought process (18–21, 25). We have previously found across MEG and fMRI studies increased medial frontal neural activity extending to cingulate activation, not only while participants self-reflect during rest states, but also during explicit reality-monitoring tasks when subjects were required to make self-agency judgments (1, 3, 4, 35). Specifically, we found increased medial frontal neural activity extending to cingulate activation during the successful encoding and memory retrieval of self-generated information, which correlated with accurate judgments of self-agency, indicating that this medial frontal site represents a neural correlate of self-agency (1, 3). We have also previously shown that this site can be directly modulated with rTMS to induce significant improvement in self-agency judgments during reality-monitoring (5).

In the present study, we now replicate and extend our prior findings (5) in a different group of participants by showing that enhancing medial SFG excitability with rTMS induced HC to significantly improve their ability to make judgments of self-agency on the reality-monitoring task. Additionally, here we implement MEG PTE metrics, measured from pre-to-post rTMS, to show that enhancing SFG excitability with rTMS not only induced increased neural information flow in the targeted SFG site, but also increased information flow from SFG to CG. Further, increased information flow from SFG to CG was observed consistently in all frequency bands. Together, these findings show that rTMS modulation of medial frontal excitability increased neural information flow from SFG to critical CG regions in the self-agency network to potentiate self-agency judgments.

We further analyzed the PTE metrics in each frequency band and found that increased information flow from SFG predicted improved self-agency judgments on the reality-monitoring task only in the HC participants who completed rTMS targeting SFG (see Figs. 3D, 4D 6C) but not in the control rTMS HC group (all p’s>.05). Specifically, increased information flow from SFG to PCL was observed in lower-frequency delta-theta and alpha bands, whereas increased information flow from SFG to CG was found in higher gamma frequencies, which predicted improved self-agency judgments on the reality-monitoring task. These results move beyond resting-state neural functional connectivity coherence-based analyses (which examine the effects of neuronal *correlations* or synchrony between regions) to a causal model showing the precise mechanisms of how enhancing SFG excitability by high-frequency rTMS modulates the temporal propagation of *directed* neural information flow from SFG to distinct regions in PCL and CG in specific frequency-bands to enhance neuronal communication that lead to the experience of self-agency.

Our results are consistent with prior studies that have shown increased neural activity in mPFC, extending to regions such as CG and PCL, in lower frequency alpha and theta bands, which is thought to reflect feedforward models of predictive coding that model the expected sensory outcome of self-generated actions to enable the initiation of self-regulated goal-directed actions (36, 37). This increased neural activity in mPFC, extending to CG and PCL, has been shown to occur consistently prior to self-generated (but not prior to externally-perceived actions), indicating these regions are important for mediating self-predictions that model the expected sensory outcome of self-generated actions, and reflect the volition in motor preparation to translate these self-predictions to initiate self-generated actions that results in the experience of self-agency (2, 9, 24–27, 36).

The sensory outcome of self-generated actions is highly predictable, compared to external actions (29–31). Thus, typically the smaller the prediction error between the actual and predicted sensory feedback, the more likely the outcome will be attributed as a self-generated (29, 31, 32). Increased gamma-band coherence has been shown to be important for neuronal communication and integration that support ‘binding’ of information during self-predictions of speech monitoring, for example, when auditory sensory feedback (i.e., what we hear when we speak) only slightly deviates from predictions of what speakers expect to hear, that leads to the experience of self-agency (34, 38, 39). We have previously found increased medial frontal activation extending to cingulate activation, shown in gamma activity, during self-predictions of speech monitoring, that results in the experience of self-agency (1, 34). Taken these findings together, this is the first multimodal study in which we combine MEG PTE metrics, from pre-to-post rTMS, to provide a causal neural basis for how enhancing medial SFG excitability improves neural information flow to CG and PCL in the self-agency network in different frequency bands to potentiate the experience of self-agency.

In summary, for the first time, we use the high spatiotemporal resolution of MEG imaging data which provide a millisecond-precision time window into the multidimensional network of neural information flow that evolve over time, space and frequency to delineate causal neurophysiological markers underlying self-agency. To-date, the neural mechanisms as to how rTMS modulates excitability of a targeted region, such as SFG, and its connected nodes in the self-agency network, remain unknown. Here, we identify the neural mechanisms of response to rTMS targeting SFG, by calculating the directed neural information flow, quantified by PTE metrics, and delineate precisely how rTMS-induced improvements in neural information flow from SFG to distinct regions in the self-agency network potentiate improvements in self-agency judgments. Such causal approaches of applying innovative MEG PTE metrics are also quick and easy to implement in clinical settings as they only require participants to complete 4min resting-state scans, measured from pre-to-post rTMS, thus facilitating the development of rapid and effective ways of measuring neural plasticity and treatment response to new neuromodulation treatment targets. In conclusion, the present research provide the first cutting-edge evidence for delineating *causal* neural mechanisms underlying self-agency and create a path towards developing new neuromodulation interventions to improve self-agency that will be particularly useful for patients with psychosis disorders who exhibit severe impairments in self-agency.

## Materials and Methods

### Participants and procedures

In the present double-blind randomized controlled trial, 30 healthy control participants (20 males, 10 females, mean age = 42.7 years, mean education = 16.8 years) volunteered to participate in the MEG study at the University of California San Francisco (UCSF) (Table 1). All methods and procedures were performed in accordance with the guidelines of the Internal Review Board (IRB) at UCSF and were approved by the IRB at UCSF. Participants were recruited through our clinicaltrial.gov site (NCT04807530) or from our previous clinical studies in schizophrenia if they had consented to be contacted for future studies. Participants were evaluated by a clinical psychologist and completed questionnaires to meet the inclusion criteria. Inclusion criteria were no Axis I or Axis II psychiatric disorder (SCID—Nonpatient edition), no current or history of substance dependence or abuse, meets MRI criteria, good general physical health, age between 18 and 64 years, right-handed, and English as the first language. Participants gave written informed consent for this protocol and then completed an MEG resting-state scan and cognitive and clinical assessments at baseline. HC were matched at a group level on age, gender, and education, and then randomly assigned to either the active 10Hz rTMS condition targeting SFG or the control 10Hz rTMS condition targeting the temporoparietal site outside the self-agency network. Immediately after the rTMS session, participants completed the MEG resting-state scan and the reality-monitoring task.

**Table 1.** Demographics (mean, SD) of Healthy Control Participants (HC)

|  | **HC in Active**<br>**rTMS to SFG** | **HC in Control**<br>**rTMS** | **p**<br>**value** |
| --- | --- | --- | --- |
| **Age (years)** | 41 (16) | 44 (18) | .60 |
| **Gender** | 10M, 5F | 10M, 5F | 1.0 |
| **Education (years)** | 17 (2.2) | 17 (1.8) | .86 |

### MRI protocol

Participants underwent T1-weighted imaging in a 3T Siemens scanner. Whole-brain structural MRI data were acquired using the following parameters (3Tesla, 3D sequence, flip angle 9°, TE 2.98 ms, TR 2300 ms, TI 900 ms, FOV 256 × 256 mm, matrix 256 × 256 × 252, NEX = 0.5, voxel dimensions 1×1×1 mm, slice thickness=1, slices per tab=160, acquisition time = 4:54 min).

### MEG data acquisition

Each participant underwent four minutes of continuous resting-state recording inside a magnetically shielded room with a 275-channel whole-head MEG system (MEG International Services Ltd., Coquitlam, British Columbia, Canada) consisting of 275 axial gradiometers. Participants were instructed to keep their eyes closed and stay awake in a supine position during the MEG scans (sampling rate = 1.2KHz). All subjects verified that they were awake throughout the resting-state scans.

To provide anatomical head models for MEG analysis, a high-resolution 3D T1-weighted whole-brain magnetic resonance imaging (MRI) was acquired for each subject using a 3T Siemens scanner. For each subject, the outline of the brain on the structural scans was extracted, and the segmented brain was treated as a volume conductor model for the source reconstruction described below. Three fiducial coils (nasion, left and right preauricular points) were placed to localize the position of the head relative to the MEG sensor array. MEG coregistration with the structural MRI for each participant was performed based on three fiducial coil positions (nasion and left and right preauricular).

### MEG data processing

The first step in data processing was to down-sample all the raw data to 1000 Hz, and remove cardiac, muscle and eye-twitch artifacts using independent component analysis. For each subject, a consecutive 4-min signal was digitally filtered using a bandpass filter of 0.7-150 Hz. Noisy channels which contained artifact due to head motion were removed based on visual inspection of the data. In addition, dual signal subspace projection (DSSP) (40) was applied to the filtered sensor signal to remove environmental noise using lead field vectors computed with an individual head model. Finally, Zapline noise filtering was applied to remove power line noise between 60 and 120 Hz (41).

After preprocessing the MEG data, source reconstruction was implemented. For source reconstruction, isotropic voxels (5 mm) were generated from a template MRI, resulting in 15,457 voxels. The generated voxels were then warped to an individual head model. For each voxel, individual magnetic lead field vectors were computed in a forward model with single-shell model approximation (42). The voxels for each participant were then indexed to 48 modules defined in the Brainnetome atlas (43). We used these 48 Brainnetome atlas modules as our ROIs.

To obtain source-localized activity, an array-gain scalar beamforming approach (44) was applied to the 4-minute resting-state sensor time series. Beamformer weights were computed in the time domain, and the data covariance matrix for beamforming was calculated for the 4-minute sensor-time series. The applied beamforming provided voxel-level source timecourses on the 5-mm volumetric grids in the brain. For each brain module from the Brainnetome atlas, the representative regional timecourse was extracted by applying principal component analysis to the voxel timecourses within each module and taking its first principal component. Before the connectivity metrics were calculated, the 48 representative timecourses were filtered into different frequency bands: delta-theta (2-8 Hz), alpha (8-12 Hz), beta (12-30 Hz), and gamma (30-50 Hz) bands. Computations of source reconstruction and connectivity metrics were performed using MATLAB.

### Phase-transfer entropy

Phase-transfer entropy (PTE) was used to evaluate pairwise directional interactions between region-of-interest (ROI) timecourses. PTE metrics compute the direction of information flow from source signal X to a target signal Y based on the phase time series (45, 46). The phase timecourses were extracted from the ROI timecourses using Hilbert transform and were used to estimate pairwise directional interactions in neural information flow during resting-state scans. The PTE from source phase timecourse X to target phase timecourse Y as derived in Engels (2017) (14) is shown below:

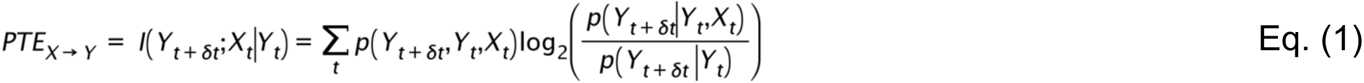

where 8t is a delay in time, p(X, Y) is a joint probability, p(X|Y) is conditional probability and I(Y;X|Z) denotes the conditional mutual information between Y and X conditioned with Z. For PTE computations between all ROIs, the source signal X represents the phase timeseries of one ROI sending phase information, and the target signal Y denotes the phase time series of another ROI receiving the phase information.

To remove bias, an estimate of PTE was also made for shuffled data which creates a null distribution, shown below:

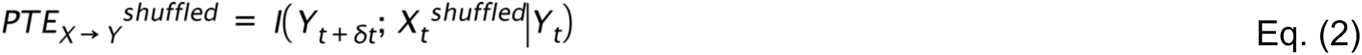

The shuffle estimate was repeated 20 times by randomizing the order of the time points for each source ROI phase time, and the shuffled PTE estimate was computed as an average of the results obtained from these 20 trials. The shuffled PTE data estimate was then subtracted from the original PTE computation to provide further statistical corrections for significance by minimizing false positives (46):

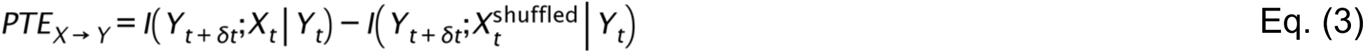

PTE for all pairwise ROIs was expressed in the form of a matrix. Since there are 48 Brainnetome regions, the dimension of the PTE matrix was 48 × 48. The regional PTEs (vectors of regional measures) can be computed by averaging values across the components of the PTE matrix along the target (*y*) array and source (*x*) array dimensions. The regional PTEs were defined as follows:

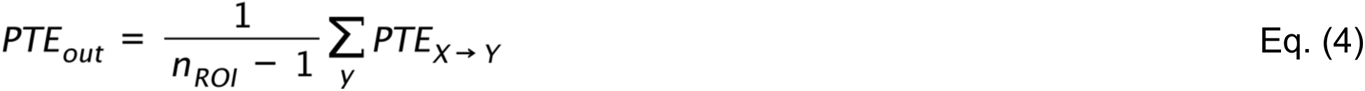

and

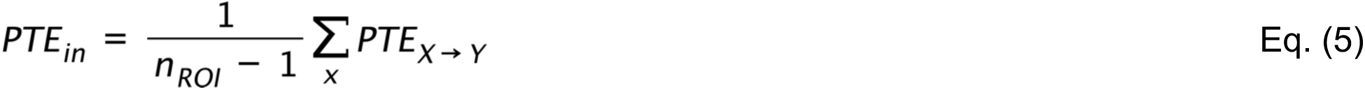

where *n_ROI_* is the number of ROIs (i.e., 48 in this case). *PTE_out_* results from averaging across the target array and indicates the regional information outflow at a source brain region; while *PTE_in_* results from averaging across the source array and denotes the regional information inflow at a target brain region.

For group-level analyses, we performed repeated-measures ANOVAs to compare changes in information flow, quantified by the PTE metrics (mbit units) after rTMS compared to baseline for each group (i.e., the active rTMS and control rTMS groups). To minimize Type I error, statistical significance was based on strict whole-brain multiple comparison FDR corrections (FDR p<.05). Regression coefficients were obtained in each group to show the strength of associations between changes in directional neural information flow (mbit units) with changes in self-agency on the reality-monitoring task (i.e., % accuracy for identifying self-generated information) after rTMS compared to baseline. We also minimized Type II errors associated with correcting for multiple regression tests, by examining regressions only in pairs that showed significant changes in neural information flow that were induced by rTMS, compared to baseline (FDR p<.05).

These MEG PTE calculations enable causal computations of changes in directed neural information flow between brain regions that is induced by rTMS, measured at the millisecond level. Using MEG PTE metrics, assayed from baseline to post-rTMS, here, we establish the precise neural mechanisms of how rTMS targeting SFG impacts the temporal propagation of neural communication in the self-agency network, compared to the control rTMS condition.

### Reality Monitoring Task

All participants completed a reality monitoring task at baseline and immediately after the rTMS session (Fig. 1). As described in previous studies (3–5, 33), the reality monitoring-task entails an encoding phase and a memory retrieval phase. All the participants completed eight runs, with 20 trials per run, totaling 160 trials for the whole task. During the encoding phase, participants were visually presented with semantically-constrained sentences with “noun-verb-noun” structures. On half the sentences, the final word was either left blank for subjects to make up on their own (i.e., *The <u>stove</u> provided the*), or was externally-given by the experimenter (i.e., *The <u>sailor</u> sailed the <u>sea</u>*). For each sentence, participants were told to pay attention to the underlined nouns for a subsequent memory test and to vocalize only the final word of each sentence. After the encoding phase, participants then completed the memory retrieval phase where they were randomly presented with the underlined noun pairs from the sentences (e.g., *sailor-sea*), and were asked to identify whether the second word was previously self-generated or externally-derived using a button box. At each time point, the sentences were completely different, containing different sets of matched semantically-constrained sentences. For each subject, accuracy for self-agency identification was computed as the percentage of correctly-identified self-generated items out of the total number of self-generated trials on the reality-monitoring task. Repeated-measures ANOVAs were implemented to examine group differences in self-agency judgments after rTMS compared to baseline.

**Fig. 1.**
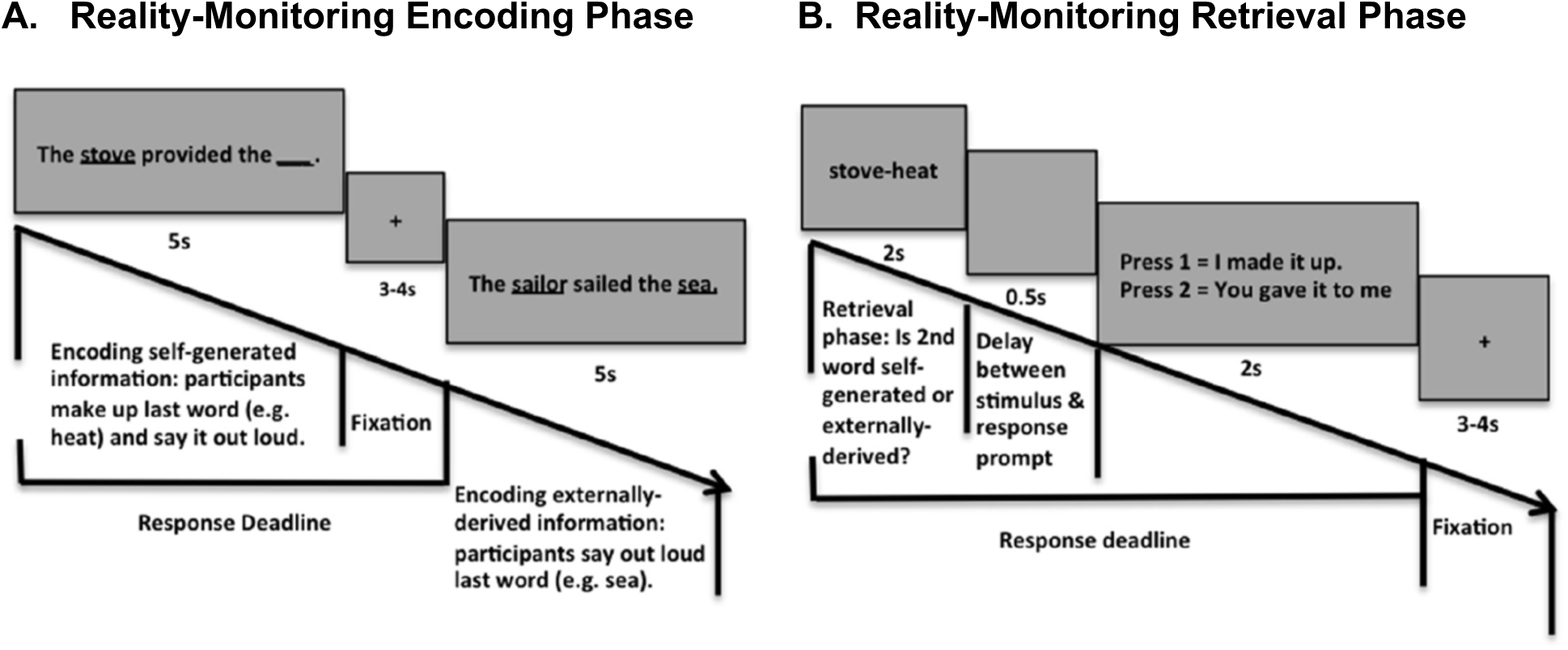
Reality Monitoring Task Design. **A.** During the encoding phase, for half the sentences, the final word was either left blank for participants to make up themselves (e.g., The stove provided the) or was externally-given by the experimenter (e.g., The <u>sailor</u> sailed the <u>sea</u>). **B.** During the retrieval phase, participants were randomly presented with the noun pairs from the sentences (e.g., stove-heat), and had to identify with a button-press whether the second word was previously self-generated or externally-derived.

### rTMS Protocol

Our TMS system is a state-of-the-art Nexstim Neuronavigated Brain Stimulator (Helsinki, Finland). This system integrates the TMS figure-8 coil with a navigational system with several landmarks on each subject’s scalp surface matched to their anatomical locations on the 3D MRI that allows for highly accurate cortical targeting individualized to each subject’s MRI anatomy. Once the targeted stimulation site is selected, the system calculates the strength of the electric field in real-time that is incident on the targeted cortical site (47, 48). The specific site that we target in subjects assigned to the active rTMS is based on our prior functional localization of medial superior frontal activity mediating self-agency (i.e., accurate identification of self-generated information) during reality monitoring across our convergent fMRI and MEG studies, labeled as the SFG ROI, on the Brainnetome atlas (3, 4) (Fig. 2A). We have previously shown that this region can be directly modulated with rTMS to induce significant improvement in self-agency abilities (5). For example, the electric field strength is shown as 65V/m in real-time that is applied to the targeted medial frontal site during rTMS in an example subject (Fig. 2A). In our previous study (5), we had established the optimal rTMS dosage parameters that maximized tolerability/comfort in the present study. The present rTMS parameters have also been deemed safe by the rTMS Consensus Guidelines. The rTMS session consisted of 120 trains of 20 pulses (2s duration of 10Hz) for 20 mins at 110% Resting Motor Threshold based on hand electromyography (5), which we and others have previously shown to maximize safety and efficacy, without a single adverse event (49–51). Due to prior reports of more discomfort and pain with higher-frequencies from our prior pilot study (5), we did not use frequencies of more than 10 Hz or theta-burst stimulation, or longer protocols (single train durations >2s).

**Fig. 2.**
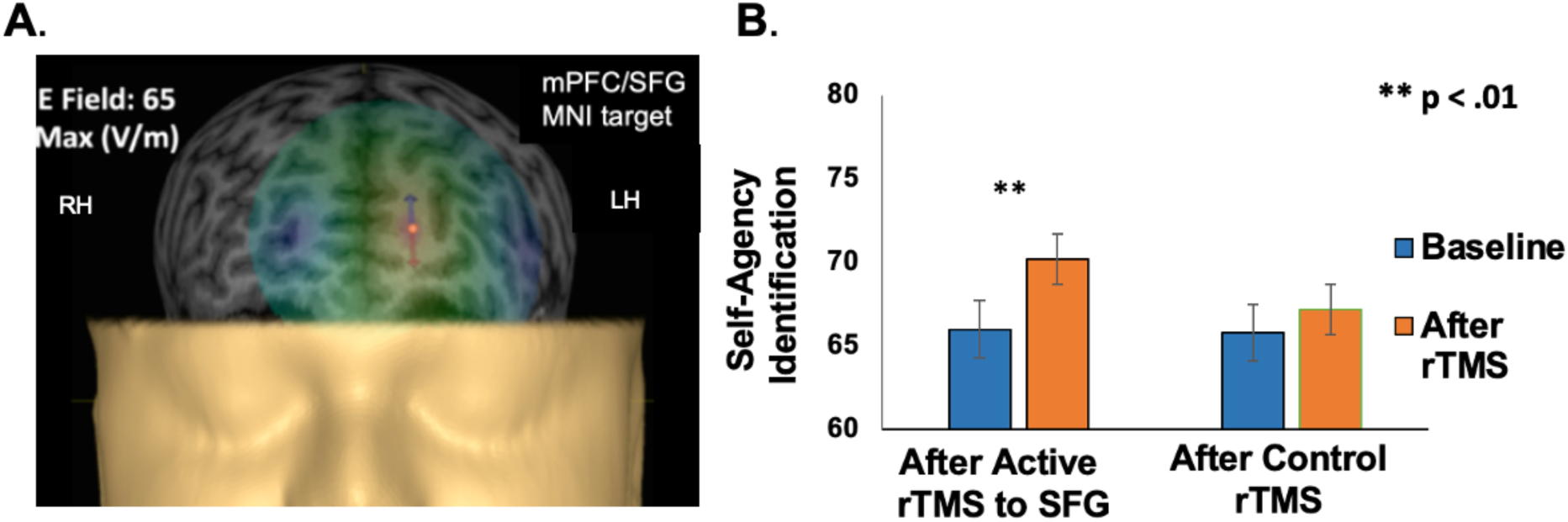
**A.** Illustration of example subject’s head model, depicting the E-field strength in real-time during active high-frequency 10 Hz rTMS to medial SFG target site (MNI:x,y,z=- 8,56,15), defined by the functional overlap of medial frontal activity underlying self-agency across our prior convergent neuroimaging fMRI and MEG studies (3, 4)**. B.** Repeated-measures ANOVA revealed HC had significant improvement in self agency judgments (i.e., % accuracy for identifying self-generated information) that was observed only after HC completed active rTMS to SFG compared to baseline, but not for HC in the control rTMS condition. *LH=left hemisphere, RH=right hemisphere

**Fig. 3.**
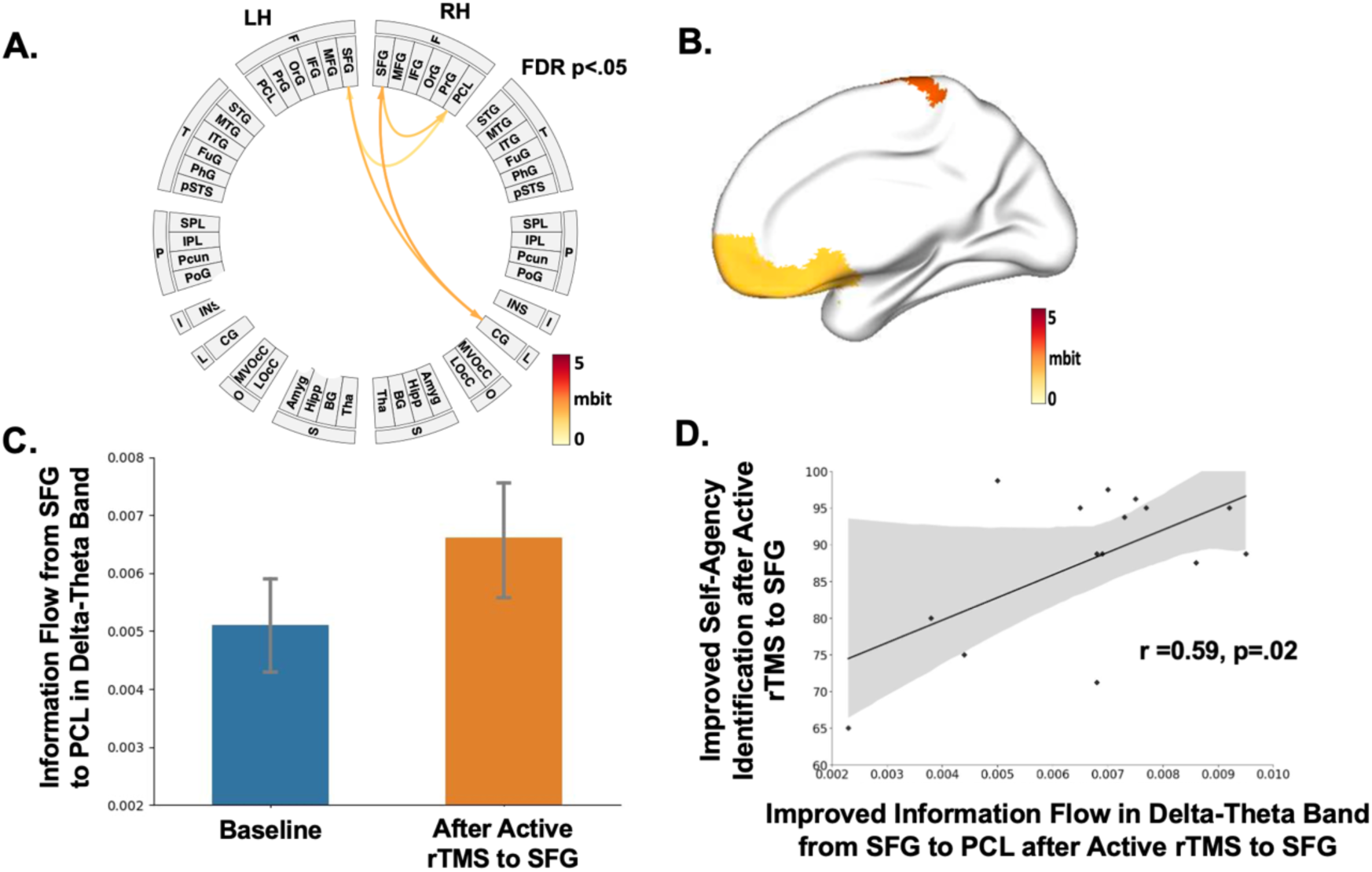
Enhancing SFG excitability by rTMS induced increased neural information flow (mbit units) from SFG to cingulate gyrus (CG) and paracentral lobule (PCL) in the self-agency network in delta-theta frequency band (2-8 Hz), compared to baseline (significant at FDR, p<.05), shown in different ways by: **(A)** Brainnetome atlas-based connectogram, **(B)** 3-D brain renderings and **(C)** histograms. **(D)** Increased information flow from SFG to PCL in delta-theta band predicted improved self-agency judgments on the reality-monitoring task (i.e., % accuracy for identifying self-generated information) after rTMS compared to baseline, only in the HC group who completed rTMS applied to SFG. *LH=left hemisphere, RH=right hemisphere

**Fig. 4.**
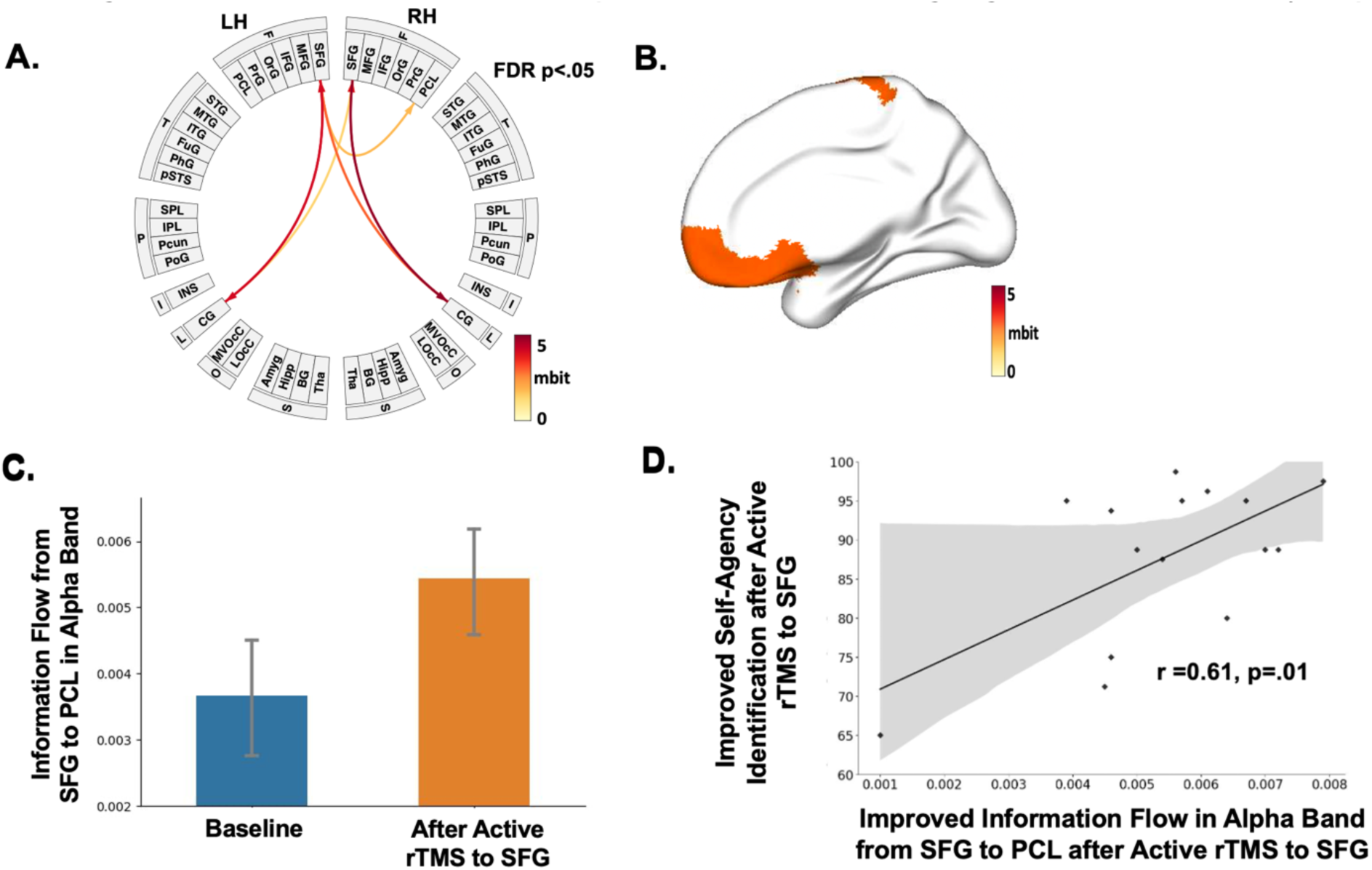
Enhancing SFG excitability by rTMS induced increased neural information flow (mbit units) from SFG to CG and PCL in the self-agency network in alpha frequency band (8-12 Hz), compared to baseline (significant at FDR, p<.05), shown by: **(A)** Brainnetome atlas-based connectogram, **(B)** 3-D brain renderings and **(C)** histograms. **(D)** Increased information flow from SFG to PCL in alpha band predicted improved self-agency judgments on the reality-monitoring task, after rTMS compared to baseline, only in the HC group who completed rTMS applied to SFG.

**Fig. 5.**
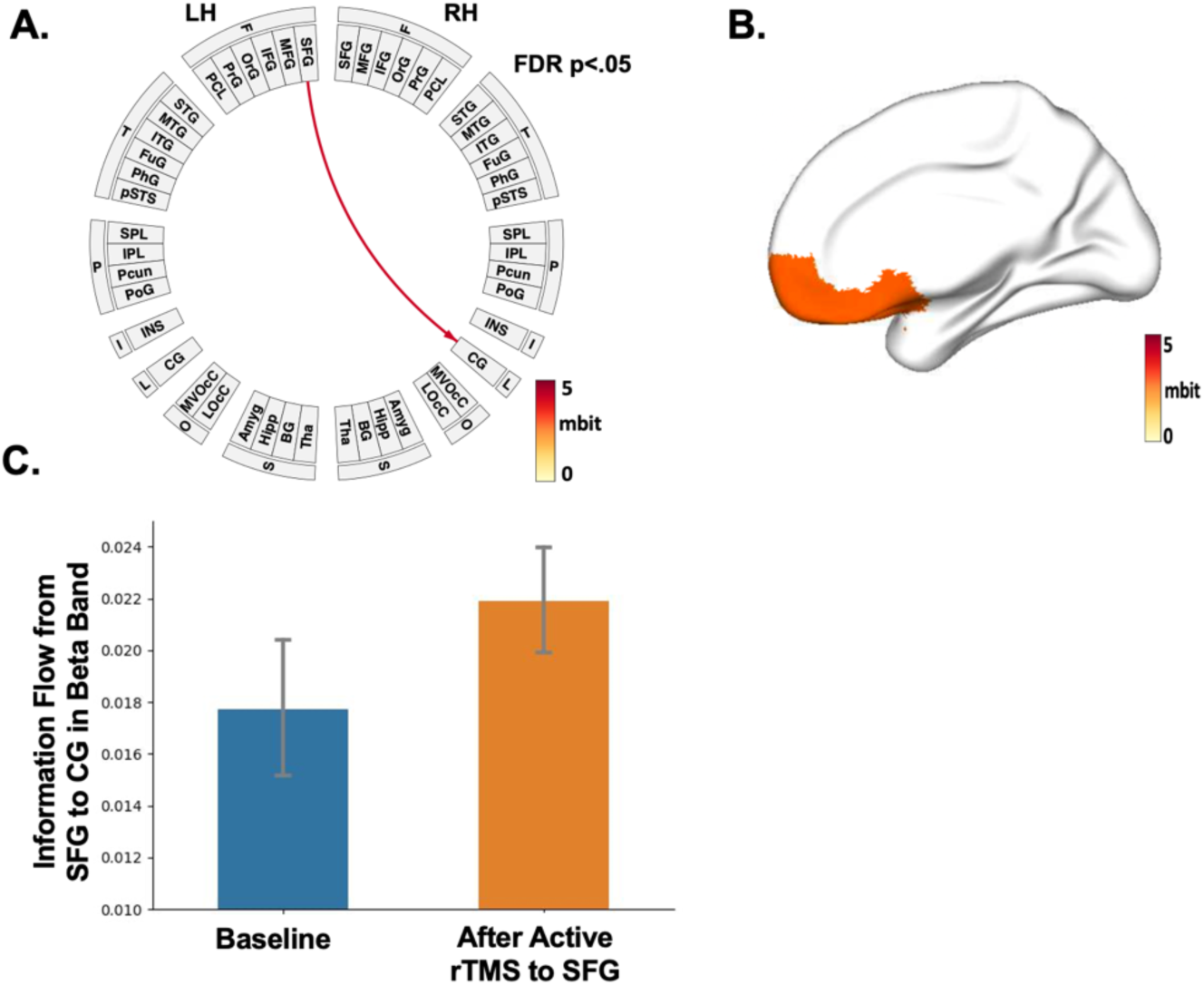
Enhancing SFG excitability by rTMS induced increased information flow (mbit units) from SFG to CG in the self-agency network in beta frequency band (12-30Hz), compared to baseline (significant at FDR, p<.05), shown by: **(A)** Brainnetome atlas-based connectogram, **(B)** 3-D brain renderings and **(C)** histograms.

**Fig. 6.**
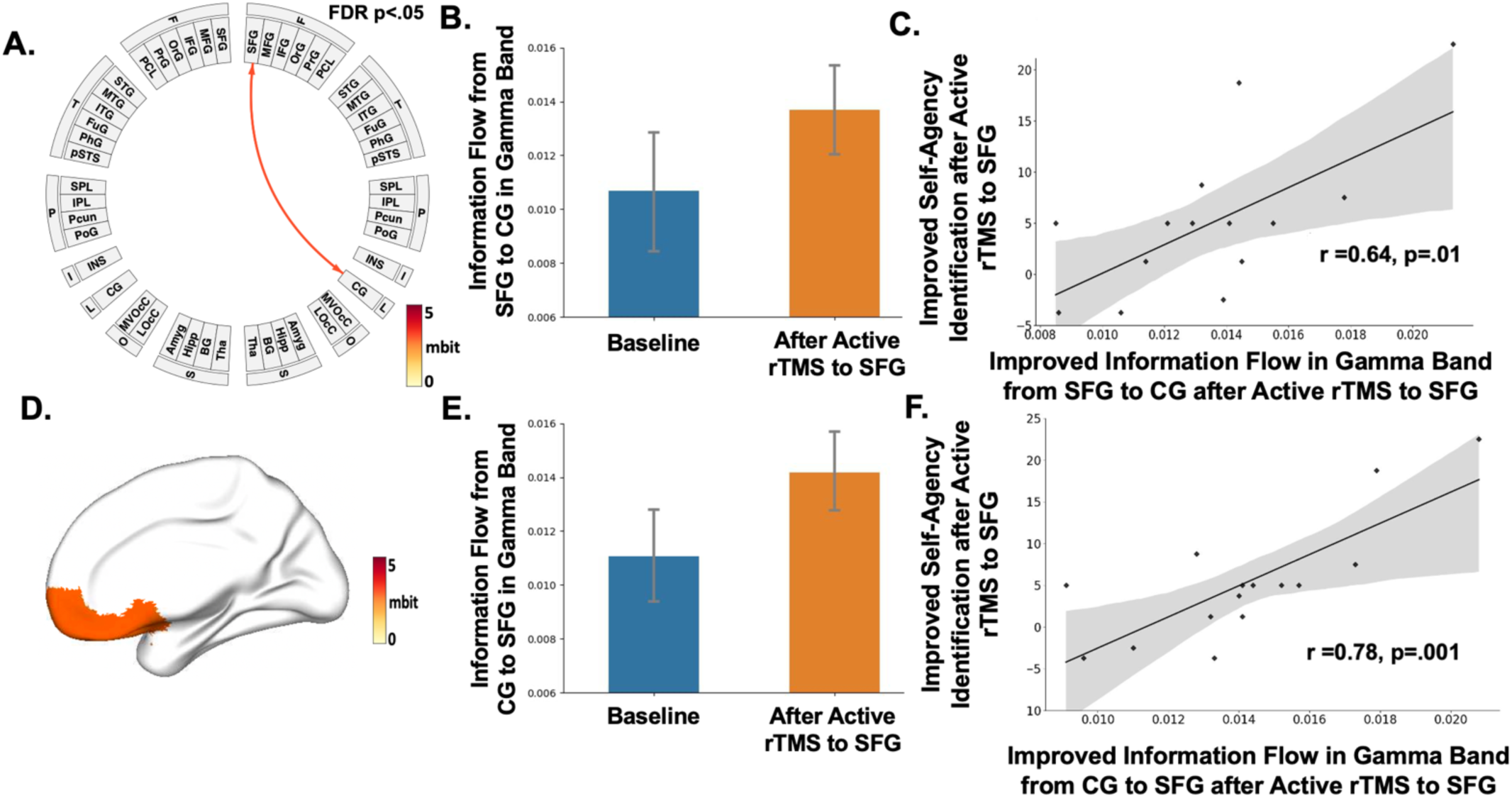
Enhancing SFG excitability by rTMS induced increased neural information flow (mbit units) from SFG to CG in the self-agency network in gamma frequency band (30-50 Hz), compared to baseline (significant at FDR, p<.05), shown by: **(A)** Brainnetome atlas-based connectogram, **(B,E)** histograms, and **(D)** 3-D brain renderings. **(C,F)** Increased information flow between SFG and CG in gamma band predicted improved self-agency judgments on the reality-monitoring task after rTMS compared to baseline, only in the HC group who completed rTMS applied to SFG.

## Acknowledgments

We thank all the participants for completing our studies. This research is supported by the Brain and Behavior Research Foundation Young Investigator Award grant (NARSAD: 28188), and an NIMH R01 grant (R01MH122897) to Karuna Subramaniam.

## Data Availability

Neuroimaging, behavioral data and code analyses will be made available from the corresponding author upon request.

---

**Supplementary Fig. 1.**
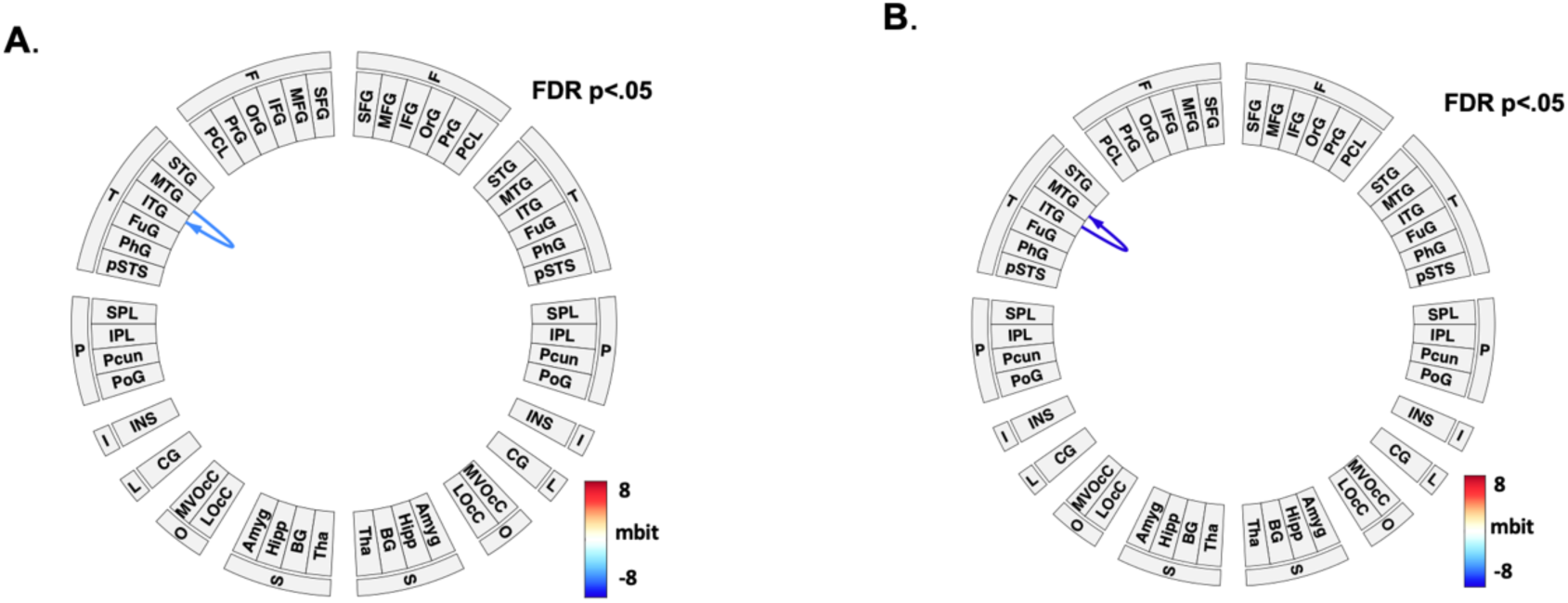
No regions revealed increased information flow after the HC group completed rTMS to the control temporoparietal site, compared to baseline. Connectograms, however, show reduced information flow between middle temporal gyrus (MTG) and inferior temporal gyrus (ITG) in **(A)** alpha band and **(B)** beta frequency bands, after HC completed control rTMS, compared to baseline (FDR, p<.05).

